# A systematic review on machine learning approaches in the diagnosis of rare genetic diseases

**DOI:** 10.1101/2023.01.30.23285203

**Authors:** P Roman-Naranjo, AM Parra-Perez, JA Lopez-Escamez

**Affiliations:** Otology and Neurotology Group CTS495, Department of Genomic Medicine, GENYO - Centre for Genomics and Oncological Research - Pfizer, University of Granada, Junta de Andalucía, PTS, Granada, Spain; Division of Otolaryngology, Department of Surgery, Instituto de Investigación Biosanitaria, ibs.GRANADA, Granada, Universidad de Granada, Granada, Spain; Sensorineural Pathology Programme, Centro de Investigación Biomédica en Red en Enfermedades Raras, CIBERER, Madrid, Spain; Meniere’s Disease Neuroscience Research Program, Faculty of Medicine & Health, School of Medical Sciences, The Kolling Institute, University of Sydney, Sydney, New South Wales, Australia

**Author notes:** **All correspondence should be addressed to:**.

**Keywords:** artificial intelligence, rare diseases, precision medicine, rare variants, DNA-sequencing, genomics

## Abstract

**Background:** The diagnosis of rare genetic diseases is often challenging due to the complexity of the genetic underpinnings of these conditions and the limited availability of diagnostic tools. Machine learning (ML) algorithms have the potential to improve the accuracy and speed of diagnosis by analyzing large amounts of genomic data and identifying complex multiallelic patterns that may be associated with specific diseases. In this systematic review, we aimed to identify the methodological trends and the ML application areas in rare genetic diseases.

**Methods:** We performed a systematic review of the literature following the PRISMA guidelines to search studies that used ML approaches to enhance the diagnosis of rare genetic diseases. Studies that used DNA-based sequencing data and a variety of ML algorithms were included, summarized, and analyzed using bibliometric methods, visualization tools, and a feature co-occurrence analysis.

**Findings:** Our search identified 22 studies that met the inclusion criteria. We found that exome sequencing was the most frequently used sequencing technology (59%), and rare neoplastic diseases were the most prevalent disease scenario (59%). In rare neoplasms, the most frequent applications of ML models were the differential diagnosis or stratification of patients (38.5%) and the identification of somatic mutations (30.8%). In other rare diseases, the most frequent goals were the prioritization of rare variants or genes (55.5%) and the identification of biallelic or digenic inheritance (33.3%). The most employed method was the random forest algorithm (54.5%). In addition, the features of the datasets needed for training these algorithms were distinctive depending on the goal pursued, including the mutational load in each gene for the differential diagnosis of patients, or the combination of genotype features and sequence-derived features (such as GC-content) for the identification of somatic mutations.

**Conclusions:** ML algorithms based on sequencing data are mainly used for the diagnosis of rare neoplastic diseases, with random forest being the most common approach. We identified key features in the datasets used for training these ML models according to the objective pursued. These features can support the development of future ML models in the diagnosis of rare genetic diseases.

## 1. Introduction

Rare diseases (RDs) continue to be a challenge to the healthcare system due to the difficulty of reaching an accurate diagnosis. Although there is no uniform international criteria, RDs are usually defined as those affecting fewer than 4-5 cases out of 10,000 individuals^1^. Considering them as a whole, RDs can be regarded as a common event, with 7,241 different RDs (http://www.orphadata.org/data/xml/en_product7.xml, updated on June 14, 2022) with an estimated accumulated prevalence of 3.5–5.9% and affecting more than 400 million people worldwide^2,3^.

Most RDs appear to be caused or modified by genetic factors; up to 80% of them are thought to have a genetic etiology^4^. Our current knowledge on this aspect is limited, existing 3,886 RDs (53.7%) linked to, at least, a gene that cause or modify the disease phenotype (http://www.orphadata.org/data/xml/en_product6.xml, updated on 14 Jun 22)^2^. The improved performance and the price reduction of Next-generation sequencing (NGS) technologies in recent years have made them more attractive for clinical applications in RDs, increasing rapidly the number of phenotype-genotype associations^5^. This has resulted in an accurate molecular diagnosis in many patients suffering from monogenic RDs, which has occasionally led to personalized treatments and improved disease management. Nevertheless, other patients with more complex disorders receive an inconclusive genetic diagnosis, placing the diagnostic yield of DNA-based NGS technologies in most studies at 40-50%^6,7^. This is mainly caused by the absence of pathogenic or likely pathogenic variants in known disease-causing genes, finding instead variants of unknown significance (VUS) or variants in novel genes not previously associated with the disease.

In this scenario of rare and complex genetic disorders where a diagnosis is not reached or a prognosis is not accurate enough, more sophisticated methods should be applied to analyze large-scale genomic data. The use of artificial intelligence (AI) and, particularly, machine learning (ML) algorithms has raised great interest in recent years due to its potential to uncover complex patterns in genomic data^8^. These ML algorithms have shown the capacity to learn from and act on large, heterogeneous datasets to extract new biological insights, improving the accuracy of the diagnosis of RDs^9–12^.

Compared to previous reviews in the field of ML and RDs, such as Schaefer *et al*.^9^ or Brasil *et al*.^13^, in this systematic review we used a different approach, investigating the role of AI/ML algorithms in the diagnosis and prognosis of RDs using genomic data. The range of options when it comes to choosing a learning algorithm or a DNA-based NGS technique to address RDs is highly variable. On the one hand, ML methods are usually divided into two main categories: supervised and unsupervised learning. Supervised ML algorithms require labeled data to solve mainly regression and classification tasks, whereas unsupervised ML algorithms address classification tasks based on unlabeled data by seeking common patterns. The review from Libbrecht *et al*. describes these algorithms in more detail and provides examples applied to genomic data^14^. On the other hand, regarding NGS techniques, there are mainly two strategies: a) to sequence the entirety of the DNA sequence (whole genome sequencing, WGS), or b) to just sequence some regions of the DNA, such as coding regions (exome sequencing, ES), or certain disease-causing genes (gene panel). Nevertheless, the raw data generated in these experiments can be processed in many ways, with different workflows depending on the aim of the study.

This systematic review presents a thorough overview of the existing evidence on the application of AI/ML algorithms to the diagnosis of RDs using DNA-based sequencing data. We conducted a comprehensive search of the literature and included studies that used a variety of ML approaches and sequencing data sources in different research settings. Our analysis focused on the evaluation of trends in the field, the ability of these approaches to identify genetic variations associated with RDs, and the potential of AI/ML to improve their diagnosis.

## 2. Methods

### 2.1. Systematic literature search and data sources

We performed a literature search using PubMed, Web of Science, and Scopus to identify relevant publications on the use of AI/ML for the diagnosis and prognosis of RDs using genomic data. We also used citation and hand searching to ensure that potentially relevant studies were retrieved. The Preferred Reporting Items for Systematic Reviews and Meta-Analyses (PRISMA) guidelines were followed to design and perform this systematic review^15^, and its protocol was registered in PROSPERO (registration number CRD42022360247).

A search in the selected databases using the search terms ‘rare AND (“artificial intelligence” OR “machine learning” OR “deep learning”) AND ((exome OR genome OR panel) AND sequencing)’ and considering publications from 2012 onward resulted in 296 abstracts. The citation and hand searching resulted in 10 additional records. The date of the last search was September 29, 2022.

The list of abstracts was screened for inclusion using the following inclusion criteria: (i) an application of AI/ML methods; (ii) a diagnostic or prognosis application using a DNA-based NGS technique (panel, exome, or genome sequencing); and (iii) an application to a RD within the orpha.net database. Non-English articles, review articles, duplicate records, and studies not relevant to any RD or AI/ML were excluded. To narrow our focus to clinical applications, we excluded animal studies as well as publications that only reported methodological aspects of AI/ML without presenting clinical data from the study population. For all articles considered relevant, the full text was reviewed using the same screening procedure as in the first stage.

### 2.2. Data extraction

All the selected articles were evaluated to gather data on five main aspects: i) study characteristics and study population (subjects included, RDs studied, study design, use of secondary data), ii) characteristics of the applied AI/ML techniques (selected ML model, programming languages used, input data, associated features, feature selection methods, model evaluation), iii) information about the DNA-based NGS technology used (type, sample collected, DNA sequencing kit, sequencing platform, read length, mean coverage), iv) the variant discovery approach (alignment method, used SNV/Indel/CNVs callers, variant annotation software, variant filtering criteria), and v) authors (number of authors and institutions involved, authors’ countries) and journal details (name, category, journal impact factor, journal citation indicator).

### 2.3. Data analysis

The data collected from selected articles were summarized and analyzed using a variety of approaches. Journal Impact Factor (JIF) scores were obtained from the Journal Citation Report (JCR) database. Bibliometric networks, including data from authors and abstracts, were constructed and visualized using VOSviewer^16^. Similarly, full-text articles were analyzed using WordStat 9.0 (Provalis Research, Montreal, Quebec, Canada) to extract main topics and keywords.

Selected articles were divided into “rare neoplastic diseases” and “other rare diseases” to enable comparisons. AI/ML models were categorized into three categories: supervised, unsupervised, and deep learning models. Input variables that the model uses to make predictions (features) were classified in 1) “clinical features”, which include information about patients’ clinical characteristics; 2) “phenotype-related features”, including data about the association between genes and phenotypes (e.g., Human Phenotype Ontology); 3) “read alignment features”, which include the properties related to read mapping and sequencing quality; 4) “genotype-related features”, including details of variants found in patients (e.g., variant allele frequency, count of variants in a certain gene, length of indel); 5) “sequence region and structural features”, including information about the region where the variant is located (e.g., gene size, GC content); 6) “network features”, which include details about the pathways in which a particular gene is involved (e.g., number of pathway, network neighbors); 7) “evolutionary/pathogenicity features”, which include pathogenicity and evolutionary conservation scores of variants (e.g., CADD, PolyPhen-2); 8) “gene expression features”, including data on gene expression; 9) “tissue-specific features”, including features which are specific for certain types of tissues; and 10) “disease-specific features”, including features which are specific for certain types of diseases. The co-occurrence of these features in the datasets used for training AI/ML models was examined and plotted using UpSetR^17^.

## 3. Results

### 3.1. Included studies

The literature search in databases identified 494 studies, with 296 remaining after removing duplicates (**Supplementary Table 1**). Among them, 93 studies were selected for full-text review, and 14 were included in the final analysis. In addition, 11 studies were identified through hand and citation searching. After screening, 8 further studies met the selection criteria of this systematic review. Thus, 22 studies were included in the final analysis (**Supplementary Table 2**). **Figure 1** shows the PRISMA flow diagram for article selection, including the reasons for excluding records.

**Figure 1:**
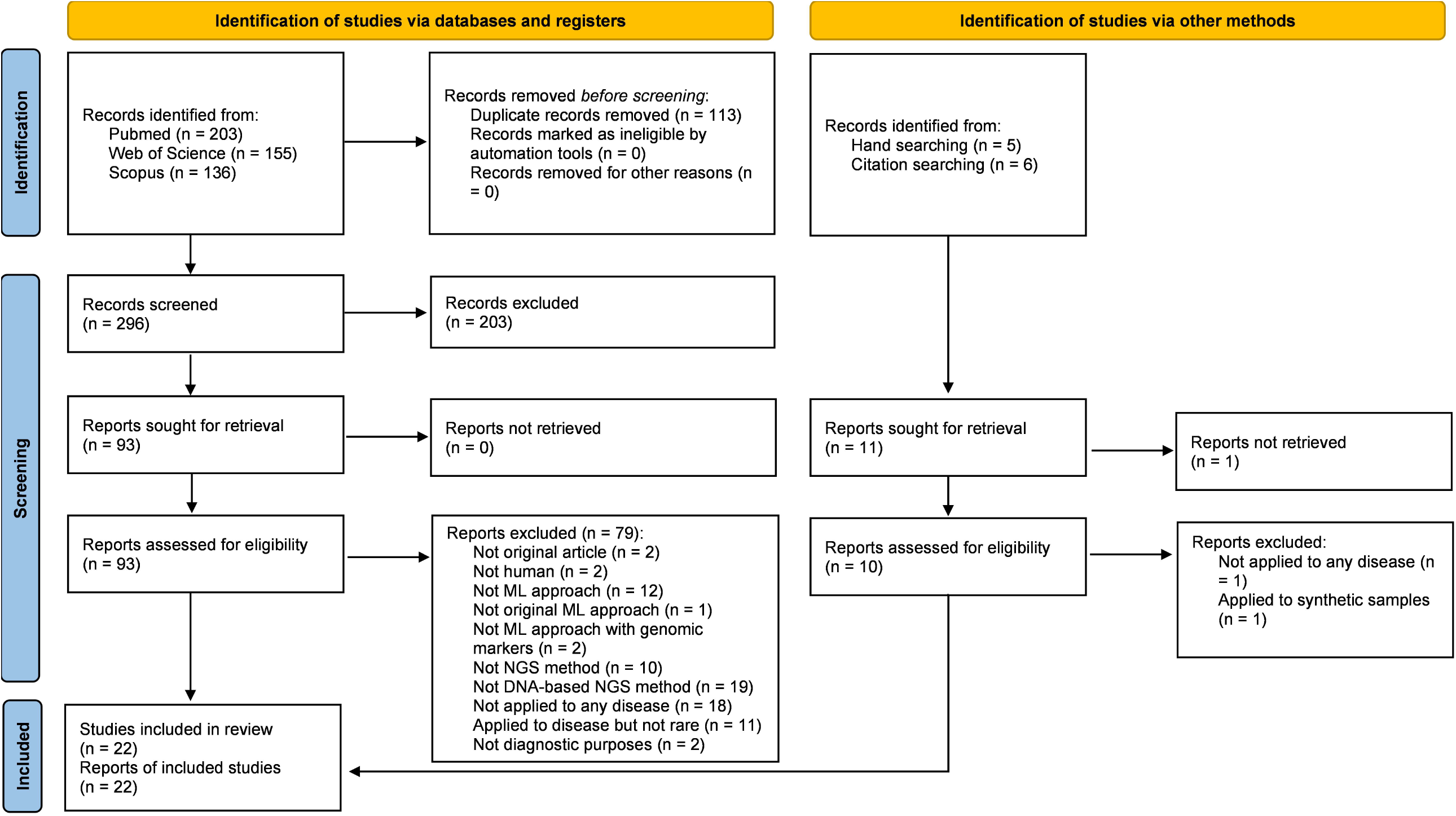
PRISMA flow diagram for the identification, screening and selection of genetic studies using AI/ML for the diagnosis of rare diseases.

### 3.2. Temporal trends and bibliometrics

To assess the temporal trends in the use of AI/ML methods for the diagnosis and prognosis of RDs using sequencing data, meta-data from included articles was retrieved (**Supplementary Table 3**). In recent years, we noticed a relative rise in the number of studies that address this challenge using AI/ML (**Figure 2A**). Most of these articles were published in journals belonging to the first quartile (90.9%) and within the “Genetics & Hereditary” JCR category (31.8%) (**Supplementary Figure 1**). It should be noted that the count for 2022 is based on studies published up to September 29, 2022.

**Figure 2:**
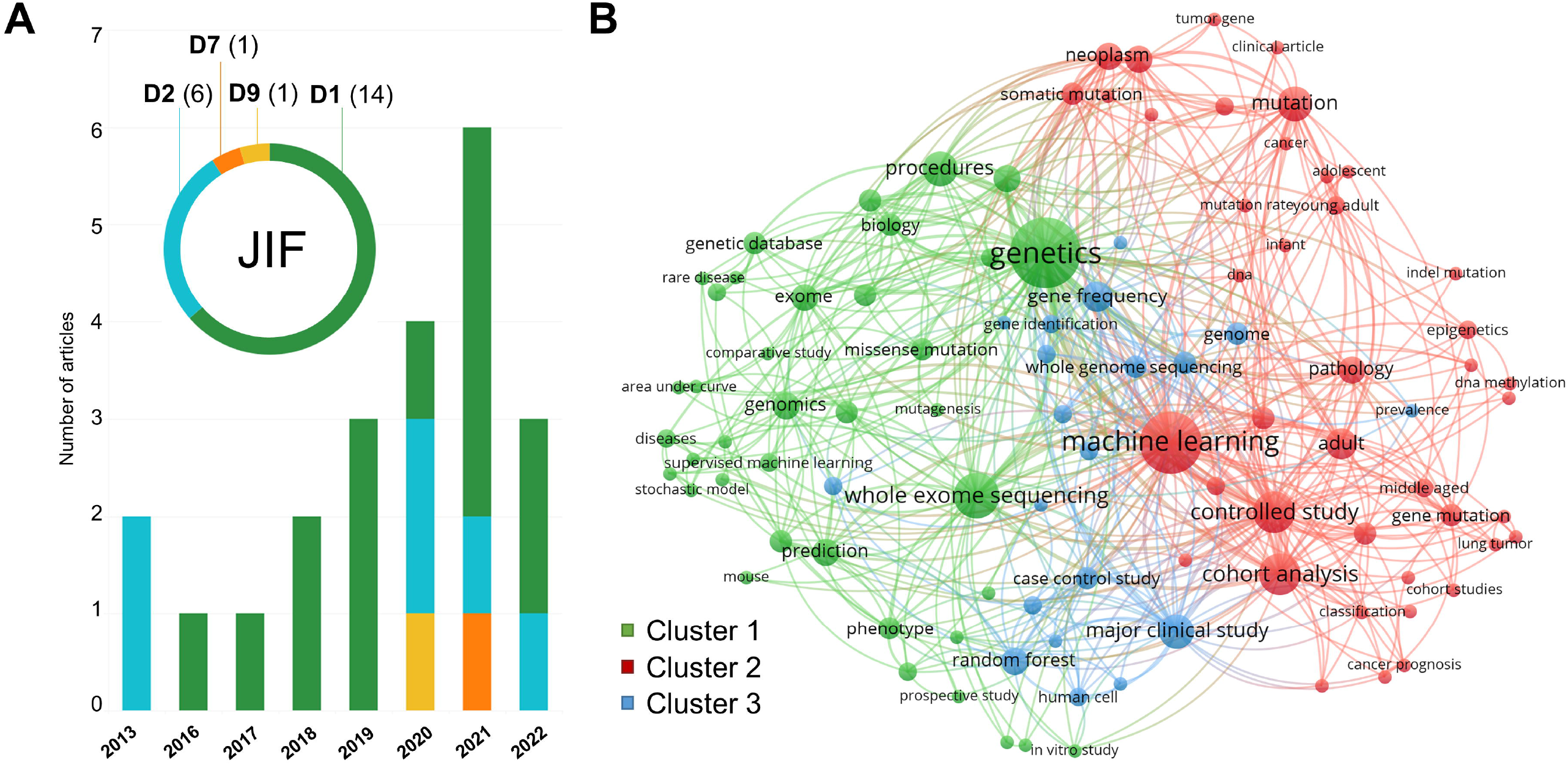
Visualization of temporal trends and bibliometrics. Panel **A)** shows the selected studies distributed per year and divided into deciles (D) according to journal impact factors (JIF). Panel **B)** displays a keyword co-occurrence network using abstracts of selected studies.

A total of 318 authors contributed to the selected articles. The bibliometric analysis showed a low level of collaboration between authors of different articles, creating 19 clusters where only 3 authors participated in 2 or more articles (**Supplementary Figure 2A**). The term co-occurrence analysis of abstracts found 100 relevant terms divided into 3 clusters that summarize the main topics of this research field. These clusters group together terms mainly associated with genetics (cluster 1), cancer (cluster 2), and methodology terms (cluster 3) (**Figure 2B**). The most frequently occurring terms in these abstracts were “genetics” (18 occurrences), “machine learning” (15 occurrences) and “whole-exome sequencing” (10 occurrences). These key terms were also among the most frequently used terms in the analysis of full-text articles, where terms such as “random forest” (59.1% of studies), “somatic mutations” (54.5% of studies), or “rare variants” (54.5% of studies) were also in a significant proportion of studies (**Supplementary Figure 2B**).

### 3.3. Application areas for AI/ML techniques

The most common disease scenario was rare neoplastic diseases (59%). The remaining studies investigated different kinds of RDs, such as developmental, neurological, or circulatory diseases **(Figure 3A)**. Exome sequencing was the most used NGS method in both rare neoplastic diseases (61.5%) and other RDs (55.5%) **(Figure 3B)**. Of note, 63.6% (14/22) of the studies employed sequencing data stored in external databases, primarily The Cancer Genome Atlas (TCGA), but also the Myocardial Genetics Consortium (MIGEN), or the Undiagnosed Diseases Network (UDN). These studies showed larger sample sizes than those using their own cohorts (**Supplementary Figure 3**), but they also showed higher intra-method variability, as seen by the mixed sample processing methods they employed (**Supplementary Figure 4**). **Supplementary Table 4** summarizes the NGS-related and sequencing data processing methods in detail.

**Figure 3:**
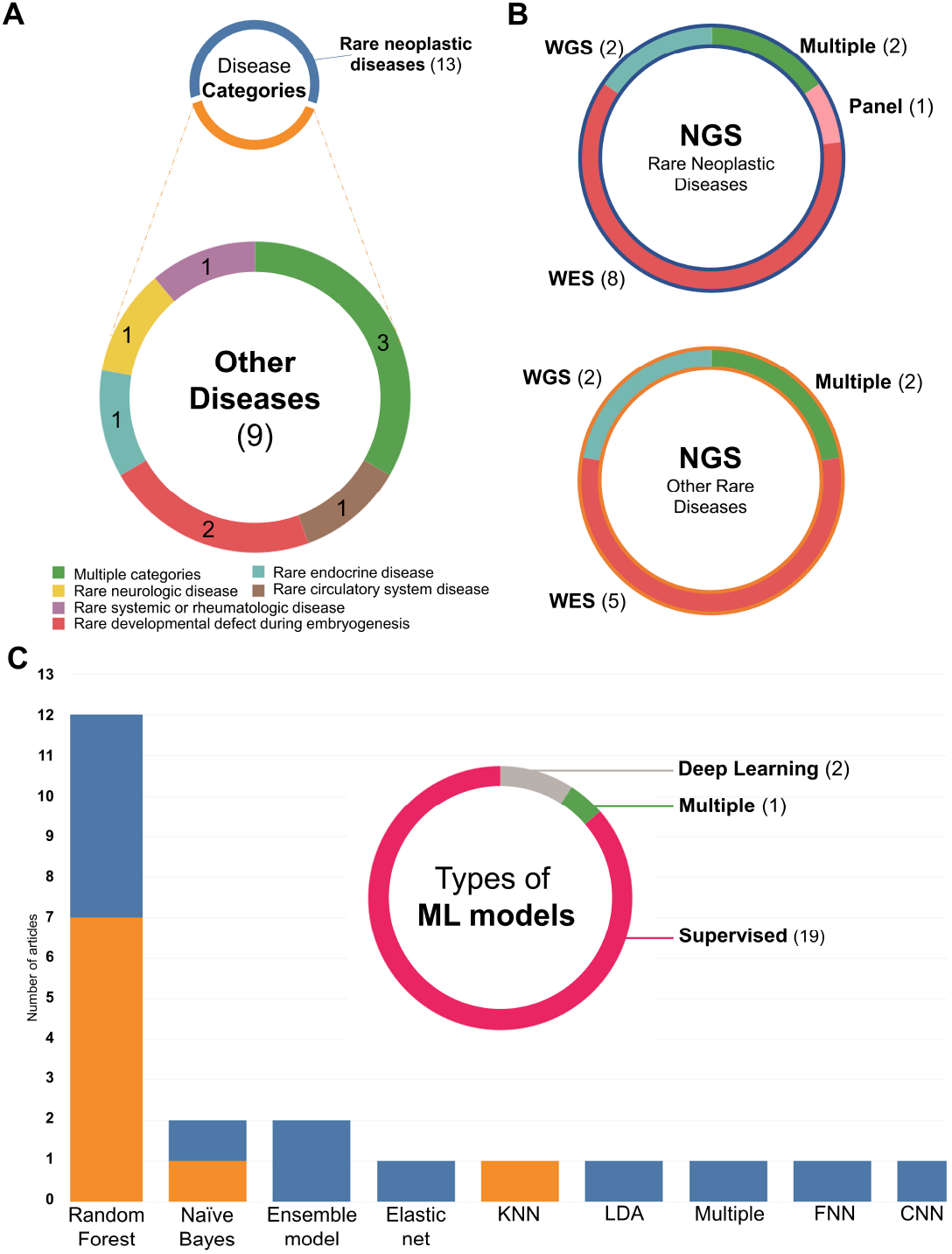
Methods and areas of application. Panel **A)** displays the distribution of rare disease identified in selected studies. Panel **B)** shows the next-generation sequencing (NGS) methods used in studies targeting rare neoplastic diseases and other rare diseases. Panel **C)** summarizes the types of machine learning algorithms applied in selected studies. Footer: KNN: K-Nearest Neighbors; LDA: Linear Discriminant Analysis; FNN: Feedforward neural network; CNN: Convolutional Neural Networks.

Supervised machine learning methods were chosen in 86.3% of the studies, with Random Forest (RF) being the most employed algorithm within this group (54.5%) **(Figure 3C)**. One study discarded the genetic features after the feature selection process, and three studies did not describe the selected features in detail, one of which was due to a commercial interest (**Supplementary Table 5**).

### 3.4. AI/ML in the study of rare genetic diseases

The objectives of AI/ML approaches in the different studies were investigated. It was found that the primary goal of using AI/ML in rare neoplastic diseases was the differential diagnosis of patients (5/13), followed by the identification of somatic mutations when a matched normal tissue was not available (4/13). In contrast, the major goals in other RDs were to prioritize variants and candidate genes (5/9) and to identify biallelic or digenic inheritance (3/9). To date, the use of AI/ML for the differential diagnosis of patients with non-neoplastic diseases is uncommon (1/9) (**Figure 4A**).

**Figure 4:**
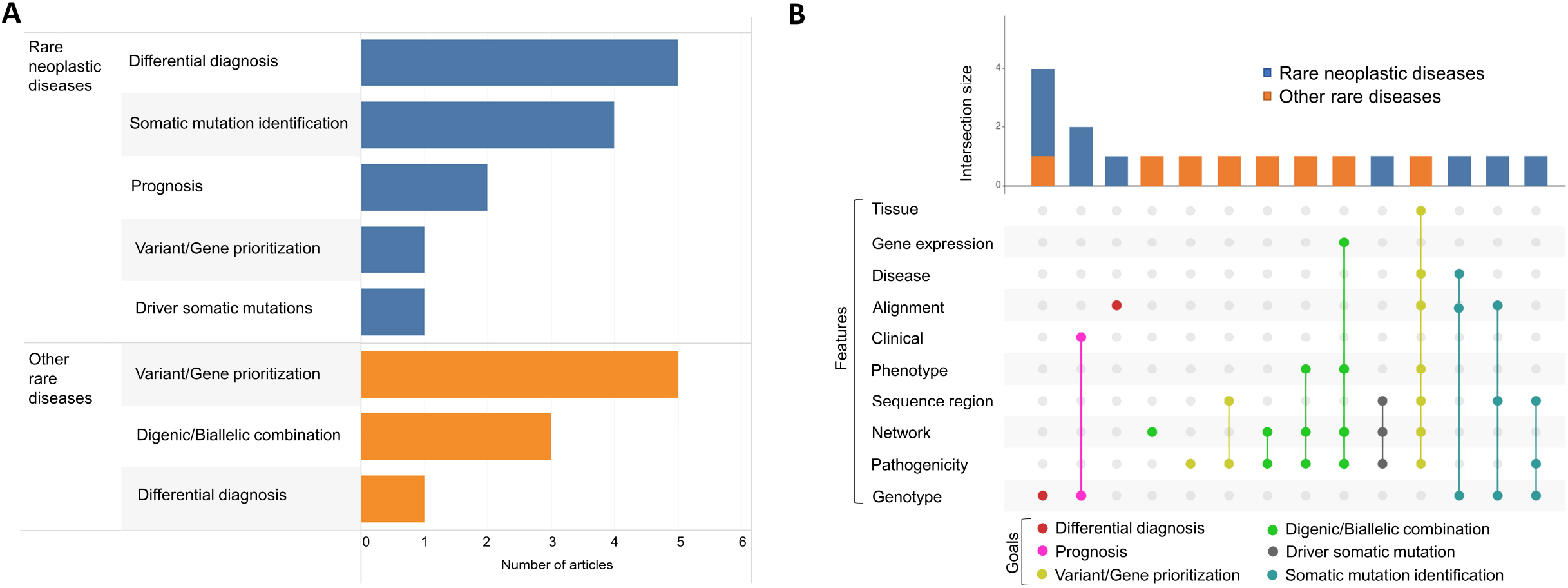
Objectives and settings of AI/ML models. Panel **A)** displays the goals of AI/ML models in rare neoplastic diseases (blue) and other rare diseases (orange). Panel **B)** contains an upset plot showing the different combinations of features in the training datasets of AI/ML models depending on the objective pursued.

Looking at the types of instances (labels) and features (attributes) of datasets used for training these AI/ML models, we found that they were distinctive and different depending on the goal pursued (**Table 1** and **Figure 4B**). For the differential diagnosis of patients, most datasets included only features related to the genotype of patients. These features primarily contained mutational load data for each gene or genomic window using collapsing methods. Models trained to predict the prognosis of RDs included clinical features (e.g., sex, age, exposure to certain substances) in addition to genotype features. The four AI/ML models aimed at finding possible pathogenic combinations of genes (digenic) or variants (biallelic) shared the usage of features related to biological networks or pathways (e.g., the associated pathway of each gene in KEGG or Reactome, network neighbors). Datasets focused on training models for variant or gene prioritization were distinguished by using features linked to predictors of variant pathogenicity at protein level and conservation across the genome of different species. Finally, for the identification of somatic mutations without a matched normal sample, the AI/ML models combined genotype features (e.g., variant allele frequency) with characteristics of the genome region where the variant is located (e.g., GC-content) or sequencing and mapping quality scores (e.g., coverage). **Supplementary Table 6** contains further information regarding the types of features mentioned above.

**Table 1:** Distinctive features identified in the datasets used for training the ML models of included studies, based on the specific goal being pursued.

| Objective AI/ML algorithm | Type of instances | Distinctive dataset feature/s | Example of feature | Use cases (ref) |
| --- | --- | --- | --- | --- |
| Stratification/Differential diagnosis | Patients | Genotype features | Burden value | [48–52] |
| Prognosis of patients | Patients | Genotype + Clinical features | Burden value + age | [53,54] |
| Variant/Gene prioritization | Genes/variants | Pathogenicity features | CADD score | [55–58] |
| Identification of digenic/biallelic combinations | Pair of genes/variants | Network features | Number of pathways shared | [59–61] |
| Identification of somatic mutations | Variants | Genotype + Sequence features | VAF + GC-content | [62–64] |

CADD: Combined Annotation Dependent Depletion; VAF: Variant allelic frequency

### 3.5. Data and code access for reproducibility

When it comes to studies that define ML models, reproducibility is a key factor. Of the selected articles, 16 studies (72.7%) provided access to the data used during the analysis; 3 studies did so only upon data request; and 3 did not explicitly declare in the text that data were available, one of which was due to commercial confidentiality. In terms of the code of AI/ML models, 16 studies (72.7%) had made it publicly available. With respect to the variant discovery approaches, all studies specified the software used for sequence alignment; 21 studies (95.5%) included information about the variant calling step; 17 studies (77.3%) did not mention the use of copy number variations (CNVs) during the analysis, and 3 studies did not state how the variants were annotated. **Supplementary Table 7** summarizes data availability and reproducibility information.

## 4. Discussion

AI/ML involve the use of algorithms to process and gain insights from data with the aim of making predictions or decisions that can be applied to a wide range of fields, including healthcare and genetics. In this systematic review, we have evaluated the latest developments in AI/ML when it comes to rare genetic conditions and examined the ways in which the use of DNA sequencing data can improve their diagnosis. In addition, we have identified some challenges and opportunities for future research in this area.

### 4.1. Exome sequencing and rare neoplastic diseases as main topics

Although to a lesser extent than in other types of diagnostic methods, such as medical imaging, AI/ML are increasingly being used in the field of RDs^9,18,19^. This trend was also found when focusing only on those studies that use DNA sequencing data to improve the diagnostic process. Through the bibliometric study carried out in this review, and the subsequent manual analyses, we found that exome sequencing was the most prevalent sequencing approach in the field, and that rare neoplastic diseases were the most prevalent clinical scenario. Exome sequencing continues to be a good starting point for the genetic diagnosis of RDs, as it provides a cost-effective and efficient way to identify disease-causing variants^20^. However, depending on the specific rare disease context, genome sequencing may be necessary to provide a complete diagnosis, including the analysis of non-coding variations, CNVs, or chromosomal rearrangements^21,22^.

Rare neoplastic diseases generally have a worse diagnosis and higher funding opportunities than other RDs, making them the type of rare disease in which AI/ML are used the most^19,23^. This is also due to the existence of public databases such as TCGA, which allow researchers to access a large amount of genomic data and use AI/ML techniques to identify patterns and make predictions^24^. When we analyzed the data on which these AI/ML models were trained, we saw that many of them (63.6%) were based on sequencing data from external sources, such as TCGA. These studies showed larger sample sizes, but also a greater diversity in sequencing technology characteristics such as read depth, different length of reads or different sequencing kits and platforms. Mixing sequencing data from different technologies, qualities, and batches can have several biases that can influence the variant calling results, affecting in turn the results of downstream analyses, and making difficult to draw accurate conclusions from the data^25^. The precision when taking clinical decisions must be maximized, so these studies must have control over these factors^26^. Different studies have shown how to approach this process^25,27^.

### 4.2. AI/ML algorithms and feature selection in genetic studies

Most of the methods utilized in the selected studies fall into the category of supervised learning (86.7%), with RF being the most common algorithm among them (73.7%). RF algorithm offers a combination of properties that makes it one of the most widely used and suitable algorithms for the study of genetic variants^28,29^. RF combines multiple decision trees (forest) that can handle high-dimensional data, capturing interactions and complex relationships between features by creating random subsets of both, data and features, at each tree. In addition, RF also allows to compute feature importances, which can be used to identify the most relevant features for the prediction task, providing interpretable models^30^. All this makes RF well suited for complex genetic problems and explains its popularity among the genetic studies.

The structure of the dataset is a fundamental and key aspect of any AI/ML model, as it is the data that the model uses to learn and make predictions. The processes of feature selection and feature engineering can have a substantial effect on the performance of the model; hence, it is essential that the final features possess relevance to the problem at hand^31^. In this systematic review, we have identified the features used by each of the selected studies and found that these features were specific to each of the objectives pursued. This insight can be valuable in understanding the current state of research in the field, and it can serve as a starting point for creating new datasets in future studies.

The results suggested that collapsing or burden methods seem to be crucial for setting up the features of datasets used to train models for the stratification or differential diagnosis of patients. These methods divide the genome into portions (bins or genes) and summarize the information contained in these segments into a burden value, which can be calculated in different ways^32,33^. This approach has shown its usefulness in finding candidate genes in different complex RDs with both genome and exome sequencing data^34–36^. Thus, applied to AI/ML tasks, this process helps to decrease the dimensionality of datasets based on genetic variants by grouping them into one value per gene or bin, which helps to reduce the curse of dimensionality and improve interpretability^37^.

On the other hand, models focused on predicting patient prognosis integrate clinical and genomic data to obtain a more complete picture of the patient and assess the risk of disease progression. Previous studies, particularly in cancer, have shown how this integration of data provides a more comprehensive and accurate assessment of patient outcome ^38,39^. Alternatively, models aimed at predicting possible pathogenic combinations of genes use features that summarize the association of these genes with the biological pathways in which they participate. The use of these features is supported by the fact that digenic diseases are usually caused by variants in genes that are functionally related and have a common pathway^40,41^.

### 4.3. Future challenges

From the results of this review, we identified some challenges that need to be addressed in future studies. When we analyzed the type of genomic data used to train the AI/ML models reviewed, we realized that most of them (77.3%) were based exclusively on single nucleotide variants or short indels, not including the analysis of CNVs. CNVs are a significant source of genetic diversity in humans that has remained understudied due to the difficulty of detection. However, today there are different algorithms for CNV detection that simplify the task considerably, as well as guidelines that help us to interpret them^42,43^. This allows the possibility of evaluating its effect on the pathogenesis and outcome of RD. On the other hand, when we examine the goals pursued in the analysis of neoplastic RDs, we can see that the differential diagnosis or stratification of patients stands out above the other objectives. This is totally different in other RDs, where, in fact, this objective is the least pursued of the 3 objectives identified, and, therefore, a field where the contribution of genetic variation to the phenotype is not well understood. The use of AI/ML algorithms on rare disease sequencing data can support the identification of novel genetic interactions, uncovering patterns and relationships that may not be immediately apparent and providing a better understanding of the regulatory mechanisms mediated by these variants in the phenotype. The use of unsupervised methods would be a possible first approach to achieve the objective of identifying clusters of patients according to their genetic background^44^.

### 4.4. Limitations

Our review is limited by the design of the systematic search and the exclusion of purely methodological articles, focusing only on those studies with clinical applications. Because of the limited number of studies available on the topic, and although it has been studied, articles have not been discarded because of the quality of the journal in which they were published (i.e., JIF), and this may have influenced, in some way, the results of this review. In addition, to reduce variability in study methodology and facilitate the analysis, we have only focused on those studies using DNA-based sequencing, not including other NGS methodologies such as RNA-seq, which are widely used in conjunction with AI/ML methodologies^45–47^.

## 5. Conclusions

We have conducted a systematic review of ML algorithms to the diagnosis of RDs using DNA-based sequencing data, providing an overview of the current state of the field and the potential of these methods to improve diagnostic accuracy. Exome sequencing is the most widely used sequencing technology and rare neoplastic diseases are the most common disease scenario. On the other hand, the goals of AI/ML algorithms in RDs using sequencing data are broad, ranging from patient stratification to the identification of possible pathogenic combinations of variants. However, we found common patterns in these goals when configuring the datasets with which these models are trained, identifying key features for each of the objectives. Finally, we identified possible future challenges, such as the use of CNV to train the AI/ML models, or the application of AI/ML for the stratification of patients with non-neoplastic RDs. Thus, this systematic review can be used as a reference for further studies, supporting the development of future ML models in the diagnosis of rare genetic diseases

## Data Availability

All data produced in the present work are contained in the manuscript.

