## Supplementary Figures for "A systematic review on machine learning approaches in the diagnosis of rare genetic diseases"

**Supplementary Figure 1**: Category and journal where selected articles were published.

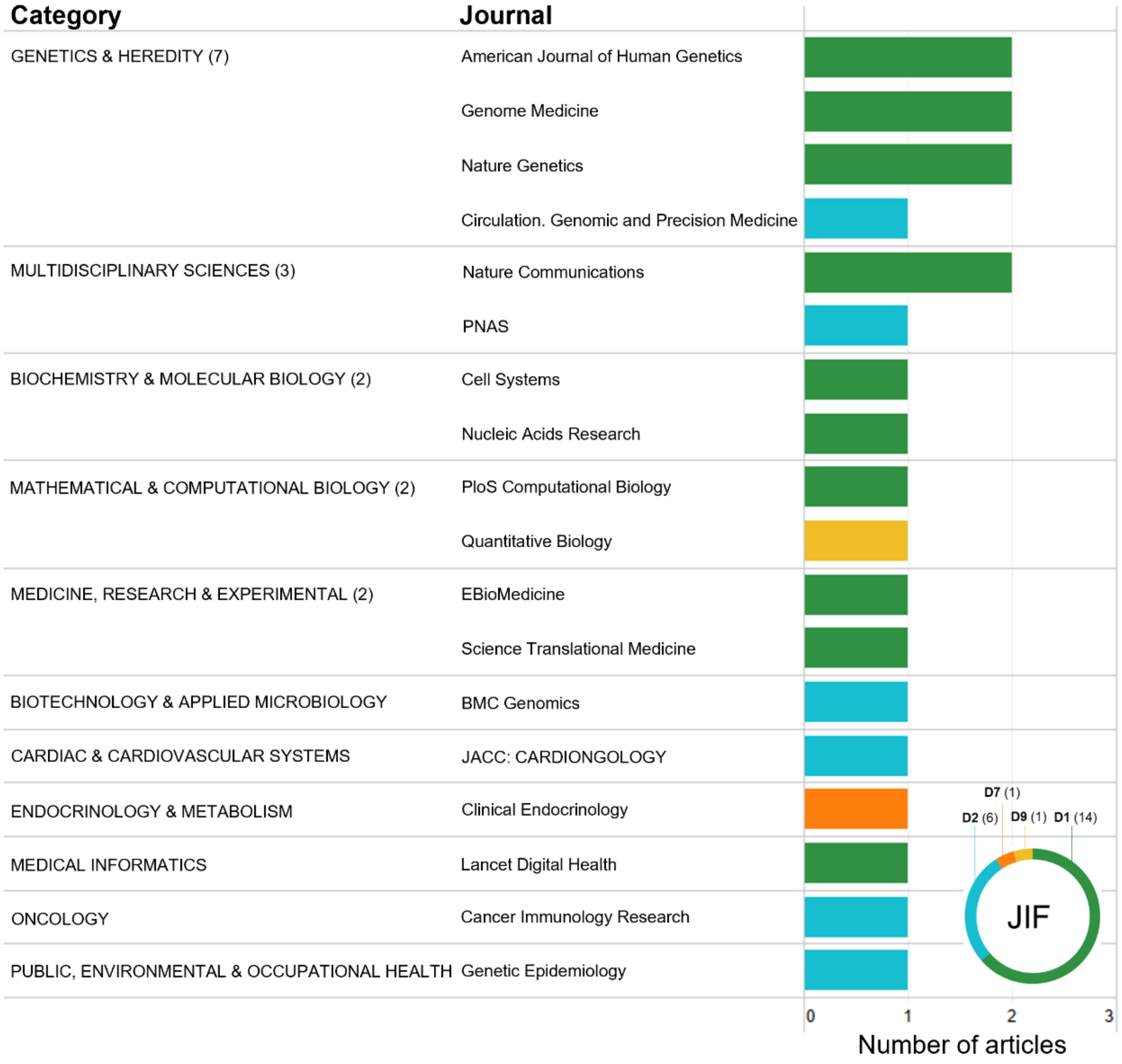


JIF: Journal Impact Factor

**Supplementary Figure 2**: A) Authors from selected articles showed a low level of collaboration, with only 3 authors participating in 2 or more articles (Rachel Karchin, Dimitrios Vitsios and Slavé Petrovski). B) Co-occurrence analysis on abstracts of selected articles.

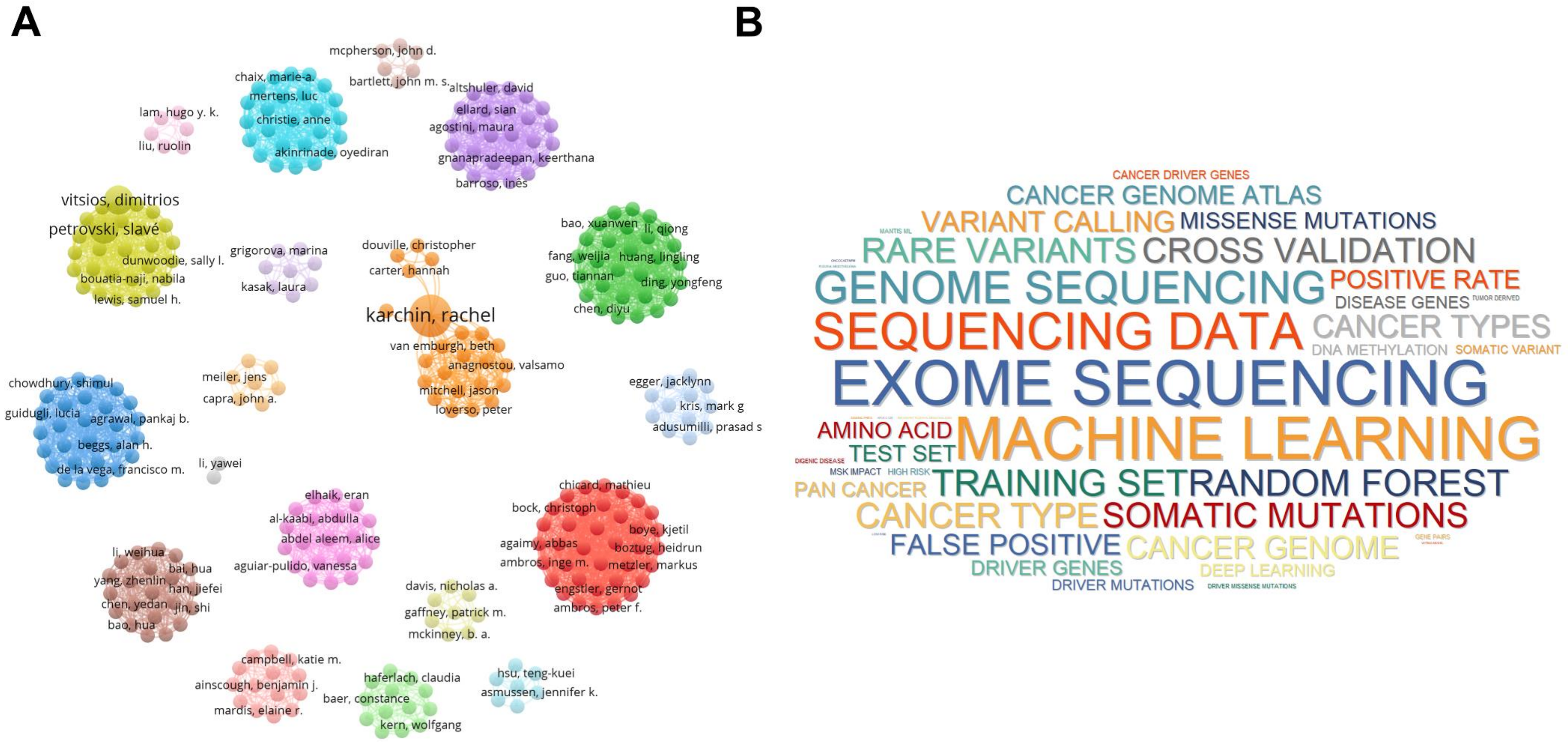

**Supplementary Figure 3**: Sample sizes of studies using A) their own cohorts and B) external cohorts.

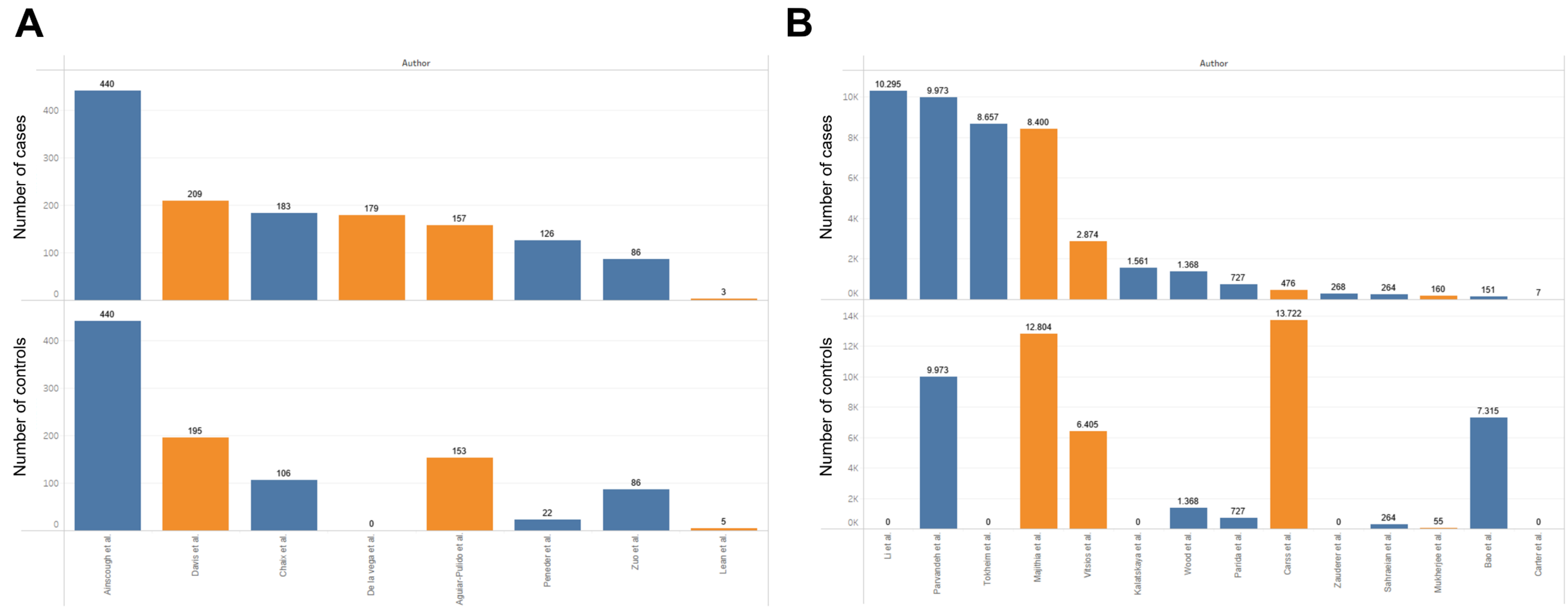

**Supplementary Figure 4:** Studies using external cohorts showed higher variability in their methods, using different sequencing platforms, read lengths and variant callers in the same study.

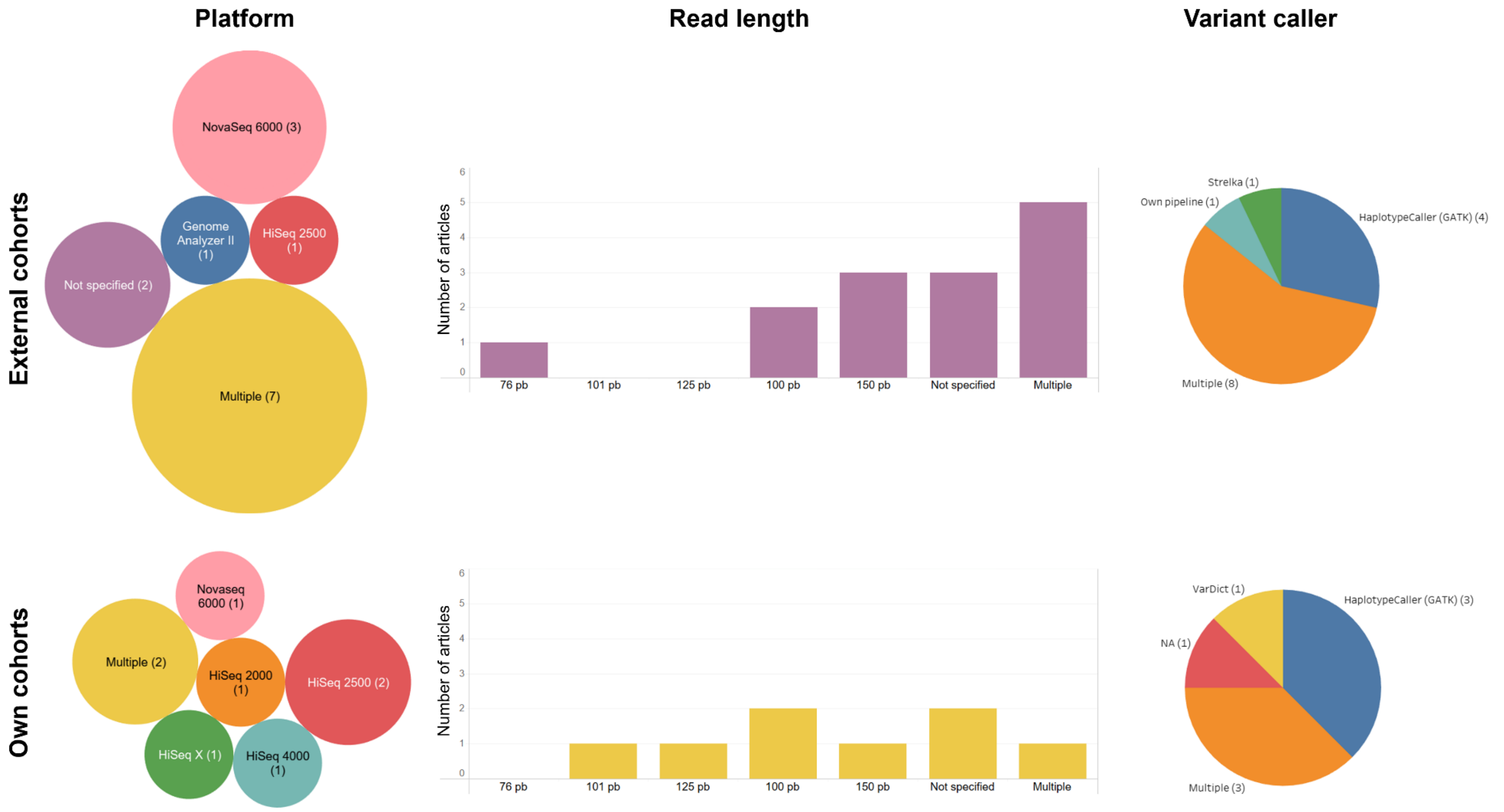
